# Trends in thiamine treatment patterns for Wernicke encephalopathy in Japan for 2010–2023: A nationwide descriptive study

**DOI:** 10.64898/2026.04.02.26350092

**Authors:** Naoki Yamagata, Yuya Kimura, Hiroki Matsui, Hideo Yasunaga

**Affiliations:** Department of Clinical Epidemiology and Health Economics, School of Public Health, The University of Tokyo, Tokyo, Japan; Department of Health Services Research, Graduate School of Medicine, University of Tokyo, Tokyo, Japan

**Author notes:** Corresponding Author: Naoki Yamagata Department of Clinical Epidemiology and Health Economics, School of Public Health, The University of Tokyo 7-3-1, Hongo, Bunkyo-ku, Tokyo 113-0033, Japan.

**Keywords:** Wernicke encephalopathy, thiamine, epidemiology, nationwide database, functional outcomes, Japan

## Abstract

**Background:** Clinical evidence on the contemporary management and functional outcomes of patients with Wernicke encephalopathy remains limited. This study aimed to clarify the nationwide patterns of thiamine administration and functional outcomes at discharge.

**Methods:** Using the Japanese nationwide inpatient Diagnosis Procedure Combination database, we identified patients hospitalized with Wernicke encephalopathy between July 2010 and March 2024. Initial intravenous thiamine doses were categorized as low (≤300 mg/day), medium (301–900 mg/day), or high (>900 mg/day). Outcomes included in-hospital mortality and functional status (Barthel Index) at discharge.

**Results:** We identified 7856 patients with Wernicke encephalopathy. Over the 13-year study period, the proportion of patients receiving initial high-dose thiamine increased markedly from 5.4% to 49.0%, while the frequency of low-dose therapy decreased from 83.0% to 37.9%. Despite prompt intervention [median time to initial administration: 0 days (interquartile range, 0 to 0 days)], 56.1% of patients were discharged with impaired activities of daily living (Barthel Index <90), and the in-hospital mortality rate was 3.8%.

**Conclusions:** High-dose thiamine treatment is increasingly implemented for Wernicke encephalopathy in Japan. Although in-hospital mortality was relatively low, the high prevalence of functional impairment at discharge, despite early treatment initiation, indicates substantial burden of Wernicke encephalopathy. Given the limited clinical evidence, further research investigating the optimal thiamine dose and develop effective primary prevention strategies for Wernicke encephalopathy is needed.

## 1. Introduction

Wernicke encephalopathy is an acute and severe neurological disorder caused by thiamine (vitamin B1) deficiency, and imposes a substantial burden on patients and healthcare systems [1]. European Federation of Neurological Societies (EFNS) guidelines (2010) and major review articles recommend initial high-dose thiamine administration (200 mg or 500 mg three times daily) for the treatment of Wernicke encephalopathy [1–3]. The recommendations for high-dose thiamine therapy are biologically plausible because experimental data suggest that cerebral thiamine requirements may not be met unless blood thiamine concentrations are raised substantially above normal [4, 5]. However, clinical evidence supporting these regimens remains limited, consisting mainly of case reports and a single randomized clinical trial with methodological limitations [1–3, 6].

Reflecting limited evidence regarding optimal treatment, a multicenter observational study conducted between 2000 and 2012 in Spain reported marked variability in initial thiamine dosing for Wernicke encephalopathy, with low-dose regimens (≤300 mg/day) being the most common [7]. However, data on thiamine dosing patterns following the 2010 guideline recommendations for high-dose therapy remain scarce and are largely limited to a few single-center studies [8, 9]. In addition to treatment patterns, evidence on the clinical outcomes of Wernicke encephalopathy remains limited. While recent large-scale studies from European countries have reported an in-hospital mortality rate of approximately 5% [7, 10, 11], data on Asian populations remain lacking. Furthermore, because Wernicke encephalopathy can severely impair functional status due to altered mental status and gait imbalance, evaluating functional outcomes, such as activities of daily living at discharge, is critically important [1]. However, data on these functional outcomes are sparse worldwide.

To address these gaps, we utilized a nationwide inpatient database in Japan to identify patients with Wernicke encephalopathy between fiscal years 2010 and 2023. Through this study, we aimed to unveil the trends in thiamine treatment patterns, as well as clinical outcomes, including in-hospital mortality and functional outcomes at discharge.

## 2. Material and Methods

### 2.1. Data Source

We analyzed data extracted from the Japanese Diagnosis Procedure Combination (DPC) inpatient database. This nationwide database consolidates data collected from more than 1000 DPC hospitals, covering approximately 50% of all acute-care inpatients in Japan [12]. The DPC includes approximately 7 million inpatient records annually. It encompasses information on patient demographics, diagnoses coded according to the International Statistical Classification of Diseases and Related Health Problems, Tenth Revision (ICD-10), medical procedures, pharmaceuticals, admission details, and discharge outcomes, including clinical measures such as the Japan Coma Scale (JCS) and the Barthel Index. Previous studies have demonstrated the high validity of primary diagnoses and procedure records in this database [13]. This study was approved by the Institutional Review Board of the University of Tokyo [approval number: 3501– (5)] and conducted in accordance with the Declaration of Helsinki. The requirement for informed consent was waived because of the anonymous nature of the database.

### 2.2. Study Design and Population

We conducted a cohort study of patients hospitalized for Wernicke encephalopathy between July 2010 and March 2024. The eligibility criteria were patient age ≥18 years and a main or admission-precipitating diagnosis of Wernicke encephalopathy (ICD-10 code: E51.2), and receipt of intravenous thiamine for at least 2 consecutive days during hospitalization. The requirement for thiamine administration was intended to enhance diagnostic specificity. For patients with multiple hospitalizations during the study period, only the first admission was included to ensure independence of observations. Additionally, based on cases identified in fiscal year 2023, we calculated the nationwide annual incidence of hospitalizations for Wernicke encephalopathy for that year. The incident cases were extrapolated to the nationwide level by adjusting for the database coverage proportion, followed by division by the annual adult population of Japan. The coverage proportion in fiscal year 2023 was derived by multiplying the proportion of DPC-database hospitals among all DPC hospitals (0.587) and the proportion of total DPC hospital beds among the total number of acute-care hospital beds in Japan (0.788) [14, 15].

### 2.3. Baseline Characteristics, Treatments, and Outcomes

We described the demographic, clinical, and hospital-related characteristics, including sex, age, body weight, body mass index, JCS score at admission, Barthel Index at admission, teaching hospital status, and annual hospital volume of Wernicke encephalopathy cases. Age was categorized into 10-year groups (18–29, 30–39, 40-49, 50-59, 60-69, 70–79, and ≥80 years). The JCS, which correlates well with the Glasgow Coma Scale [16, 17], was categorized as follows: 0 (alert), 1–3 (confused), 10–30 (somnolent), and 100–300 (comatose). Barthel Index scores were classified as follows: 100 (independent), 90–95 (slightly dependent), 65–85 (moderately dependent), and 0–60 (severely dependent).

Regarding intravenous thiamine treatment patterns, we assessed the time interval from admission to the first administration, the dose administered on the second day of therapy, and the total duration of treatment. The dose administered on the second day of therapy was considered to represent the intended initial thiamine dose rather than the first-day dose. This is because intravenous thiamine is typically administered two or three times daily owing to its short half-life [18], but since the database only records the total daily dose, using the first-day dose could underestimate the intended initial dose; for example, a patient admitted at night may receive treatment only once on the day of admission. The second-day dose was categorized as low (≤300 mg/day), medium (301–900 mg/day), or high (>900 mg/day) for descriptive analyses. In addition, the distribution of the second-day dose was visualized using a histogram. Temporal trends in the annual proportion of the second-day dose categories (low, medium, high) were illustrated in a figure and evaluated independently using the Cochran–Armitage trend test. We also identified other drugs used during hospitalization besides thiamine that may have been used to treat Wernicke encephalopathy or related comorbid conditions. These drugs included folic acid, methylcobalamin (vitamin B12), magnesium, antipsychotics, oral benzodiazepines, and anti-seizure drugs [7, 19, 20]. Antipsychotics included typical and atypical agents in oral and intramuscular formulations. Oral benzodiazepines included agents with any duration of action. Anti-seizure drugs included intravenous benzodiazepines, propofol, fosphenytoin, phenytoin, phenobarbital, thiopental, valproic acid, carbamazepine, lamotrigine, levetiracetam, and lacosamide.

The outcomes included in-hospital mortality, admission to the intensive care unit, readmission within 30 days of discharge, JCS score at discharge, Barthel Index at discharge, length of hospitalization, and total healthcare costs. Healthcare costs were converted to US dollars (USD) based on the average annual exchange rate of Japanese yen to USD for each study year.

### 2.4. Statistical Analysis

Continuous variables are presented as medians and interquartile ranges (IQR), whereas categorical variables are presented as frequencies and percentages. A two-sided P value of <0.05 was considered statistically significant. Data analyses were performed using Microsoft SQL server Management Studio version 14 and R version 3.6.3 (R Foundation for Statistical Computing, Vienna, Austria).

### 2.5. Exploratory Analysis

We conducted an exploratory analysis to investigate differences in baseline characteristics, treatment patterns, and clinical outcomes according to the second-day thiamine dose category (low, medium, high). To assess differences across groups, we calculated the maximum pairwise absolute standardized difference (ASD) for each category [21]. These differences were presented as a plot of maximum ASDs, with a value >0.1 denoting a notable difference [22]. These exploratory analyses were intended to describe the baseline characteristics, treatment practices, and clinical outcomes, rather than to establish causal inference.

## 3. Results

### 3.1. Study Population

During the study period, 9812 admissions with a diagnosis of Wernicke encephalopathy were identified. After excluding 325 repeat admissions, 9487 patients remained. We also excluded 37 patients aged <18 years and 1594 patients who did not receive intravenous thiamine for at least 2 consecutive days. Consequently, 7856 patients were included in the analysis. The number of hospitalized patients in fiscal year 2023 was 641, corresponding to an estimated annual incidence of 1.28 per 100,000 person-years.

### 3.2. Demographic and Clinical Characteristics

Among the 7856 patients, the median age was 65 years (IQR, 56 to 74 years), 22.4% were women, and the median body mass index was 19.5 kg/m^2^ (IQR, 17.2 to 22.1 kg/m^2^) (Table 1). Patients aged 60–69 years accounted for the largest proportion of the cohort. Among patients with available data at admission, 69.4% had altered mental status (JCS score ≥1) and 83.1% had a Barthel Index <90.

**Table 1.**
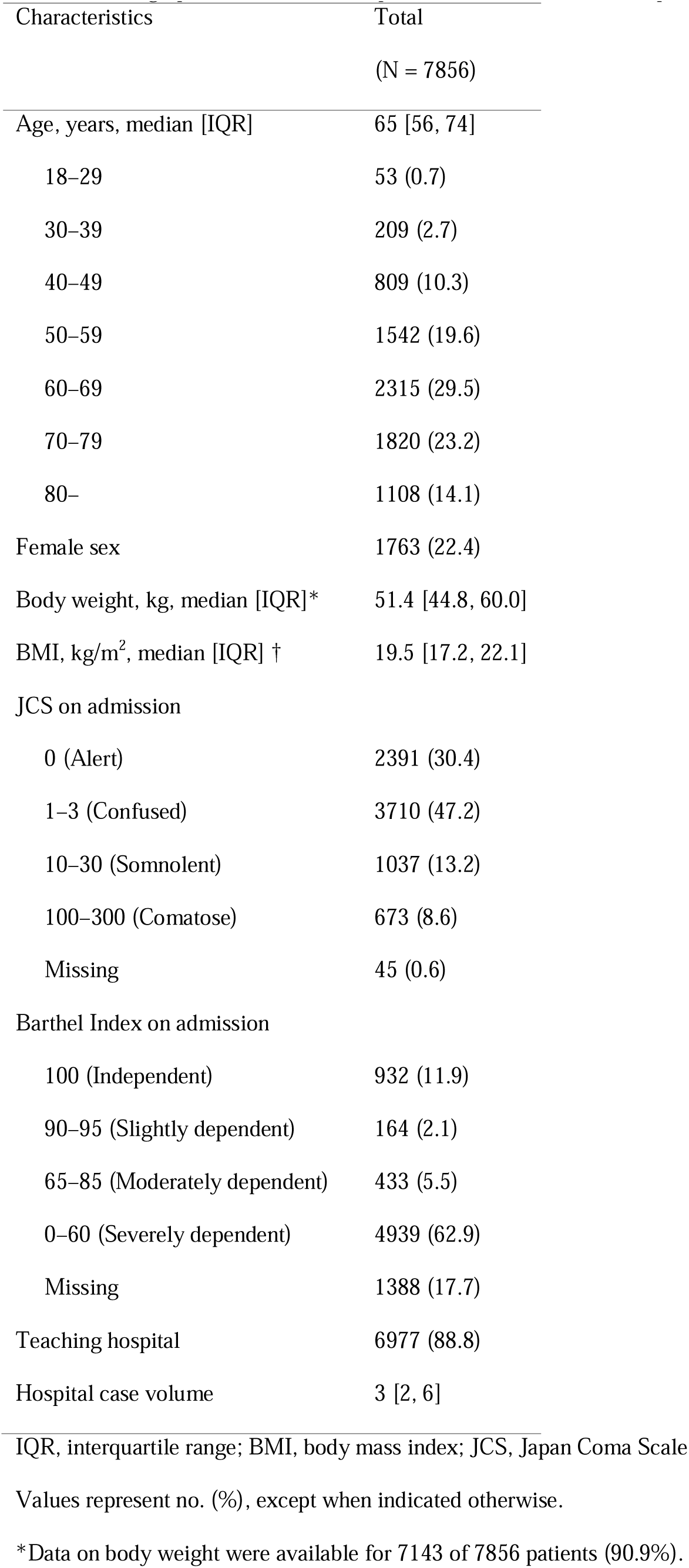
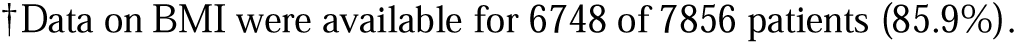
Demographic, clinical, and hospital-related characteristics of patients with Wernicke encephalopathy.

| Characteristics | Total |
| --- | --- |
|  | (N = 7856) |
| Age, years, median [IQR] | 65 [56, 74] |
| 18–29 | 53 (0.7) |
| 30–39 | 209 (2.7) |
| 40–49 | 809 (10.3) |
| 50–59 | 1542 (19.6) |
| 60–69 | 2315 (29.5) |
| 70–79 | 1820 (23.2) |
| 80– | 1108 (14.1) |
| Female sex | 1763 (22.4) |
| Body weight, kg, median [IQR]* | 51.4 [44.8, 60.0] |
| BMI, kg/m<sup>2</sup>, median [IQR] † | 19.5 [17.2, 22.1] |
| JCS on admission |  |
| 0 (Alert) | 2391 (30.4) |
| 1–3 (Confused) | 3710 (47.2) |
| 10–30 (Somnolent) | 1037 (13.2) |
| 100–300 (Comatose) | 673 (8.6) |
| Missing | 45 (0.6) |
| Barthel Index on admission |  |
| 100 (Independent) | 932 (11.9) |
| 90–95 (Slightly dependent) | 164 (2.1) |
| 65–85 (Moderately dependent) | 433 (5.5) |
| 0–60 (Severely dependent) | 4939 (62.9) |
| Missing | 1388 (17.7) |
| Teaching hospital | 6977 (88.8) |
| Hospital case volume | 3 [2, 6] |

IQR, interquartile range; BMI, body mass index; JCS, Japan Coma Scale
Values represent no. (%), except when indicated otherwise.
\*Data on body weight were available for 7143 of 7856 patients (90.9%).
†Data on BMI were available for 6748 of 7856 patients (85.9%).

### 3.3. Treatments

The median time interval from admission to the first administration of intravenous thiamine was 0 days (IQR, 0 to 0 days), and the median total duration was 7 days (IQR, 4 to 9 days) (Table 2). The median second-day dose of thiamine was 250 mg/day (IQR, 100 to 1300 mg/day), and 55.1%, 13.9%, and 31.0% of patients were classified into the low-, medium-, and high-dose categories, respectively. The histogram showed bimodal peaks at doses of 100 mg/day and 1500 mg/day (Fig. 1a). The proportion of patients in the high-dose group increased from 5.4% to 49.0% (p<0.001) between fiscal years 2010 and 2023, while the proportion in the low-dose group decreased from 83.0% to 37.9% (p<0.001) between fiscal years 2010 and 2023 (Fig. 1b). In contrast, the proportion of patients in the medium-dose group increased from 11.7% to 13.1%, but the change was not statistically significant (p=0.14).

**Fig. 1.**
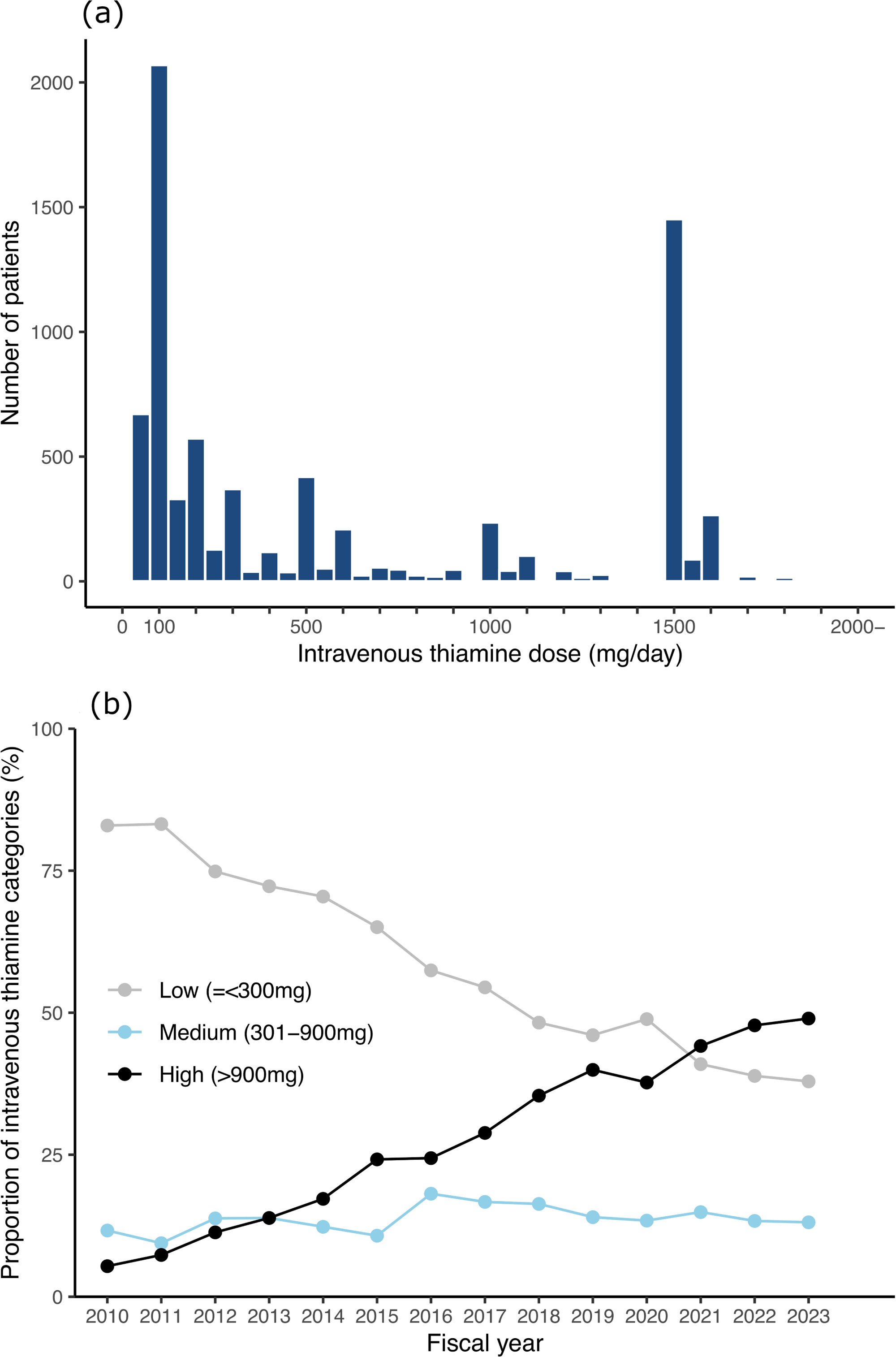
Second-day intravenous thiamine dose and temporal trends in patients with Wernicke encephalopathy. (a) Histogram of the second-day dose distribution (b) Temporal trends in the three dose categories from fiscal years 2010 to 2023: low (≤300 mg/day), medium (301–900 mg), and high (>900 mg)

**Table 2.** Treatment patterns in patients with Wernicke encephalopathy.

| Treatment patterns | Total |
| --- | --- |
|  | (N = 7856) |
| Intravenous thiamine |  |
| Time from admission to therapy, days, median [IQR] | 0 [0, 0] |
| Duration, days, median [IQR] | 7 [4, 9] |
| Dose on the second administration day, mg/day, median [IQR] | 250 [100, 1300] |
| Low ($\leq 300$ mg) | 4329 (55.1) |
| Medium (301–900 mg) | 1095 (13.9) |
| High ($> 900$ mg) | 2432 (31.0) |
| Folic acid† | 2002 (25.5) |
| Methylcobalamin (vitamin B12) * | 3545 (45.1) |
| Magnesium* | 2732 (34.8) |
| Antipsychotics* | 3104 (39.5) |
| Oral benzodiazepine* | 3593 (45.7) |
| Anti-seizure drugs* | 2424 (30.9) |

IQR, interquartile range
Values represent no. (%), except when indicated.
\*Prescribed at least once during hospitalization.

### 3.4. Outcomes

The rates of in-hospital mortality, intensive care unit admission, and readmission within 30 days of discharge were 3.8%, 8.8%, and 3.3%, respectively (Table 3). The median length of hospitalization was 23 days (IQR, 12 to 41 days). At discharge, 30.4% of patients with available JCS scores (n = 7539) had altered mental status (JCS score ≥1), and 56.1% of patients with available Barthel Index values (n = 6755) had scores <90.

**Table 3.** Outcomes of patients with Wernicke encephalopathy.

| Outcomes | Total |
| --- | --- |
|  | (N = 7856) |
| In-hospital mortality | 299 (3.8) |
| Admission to the intensive care unit | 695 (8.8) |
| Readmission within 30 days of discharge | 260 (3.3) |
| Total healthcare costs, USD, median [IQR] | 8500 [4920, 13826] |
| Length of hospitalization, days, median [IQR] | 23 [12, 41] |
| JCS at discharge |  |
| 0 (Alert) | 5247 (66.8) |
| 1–3 (Confused) | 2144 (27.3) |
| 10–30 (Somnolent) | 105 (1.3) |
| 100–300 (Comatose) | 43 (0.5) |
| Missing | 317 (4.0) |
| Barthel Index at discharge |  |
| 100 (Independent) | 2547 (32.4) |
| 90–95 (Slightly dependent) | 420 (5.3) |
| 65–85 (Moderately dependent) | 809 (10.3) |
| 0–60 (Severely dependent) | 2979 (37.9) |
| Missing | 1101 (14.0) |

IQR, interquartile range; JCS, Japan Coma Scale
Values represent no. (%), except when indicated otherwise.

### 3.5. Exploratory Analysis

Comparisons across the second-day intravenous thiamine dose (low, medium, high) categories are shown in Table 4 and Fig. 2. Patients who received higher thiamine doses had a higher JCS score on admission (max ASD = 0.19) and were more likely to be treated at teaching hospitals (max ASD = 0.36) and at hospitals with higher Wernicke encephalopathy case volumes (max ASD = 0.54). In addition, higher thiamine doses were associated with earlier thiamine administration (max ASD = 0.17) and more frequent use of folic acid supplementation (max ASD = 0.18), magnesium (max ASD = 0.45), antipsychotics (max ASD = 0.17), oral benzodiazepines (max ASD = 0.21), and anti-seizure drugs (max ASD = 0.16). Clinical outcomes such as in-hospital mortality, readmission within 30 days of discharge, and total healthcare costs exhibited no notable differences, although the high-dose group had more frequent intensive care unit admission (max ASD = 0.12), longer length of hospitalization (max ASD = 0.12), and worse JCS (max ASD = 0.16) and Barthel Index scores (max ASD = 0.11).

**Fig. 2.**
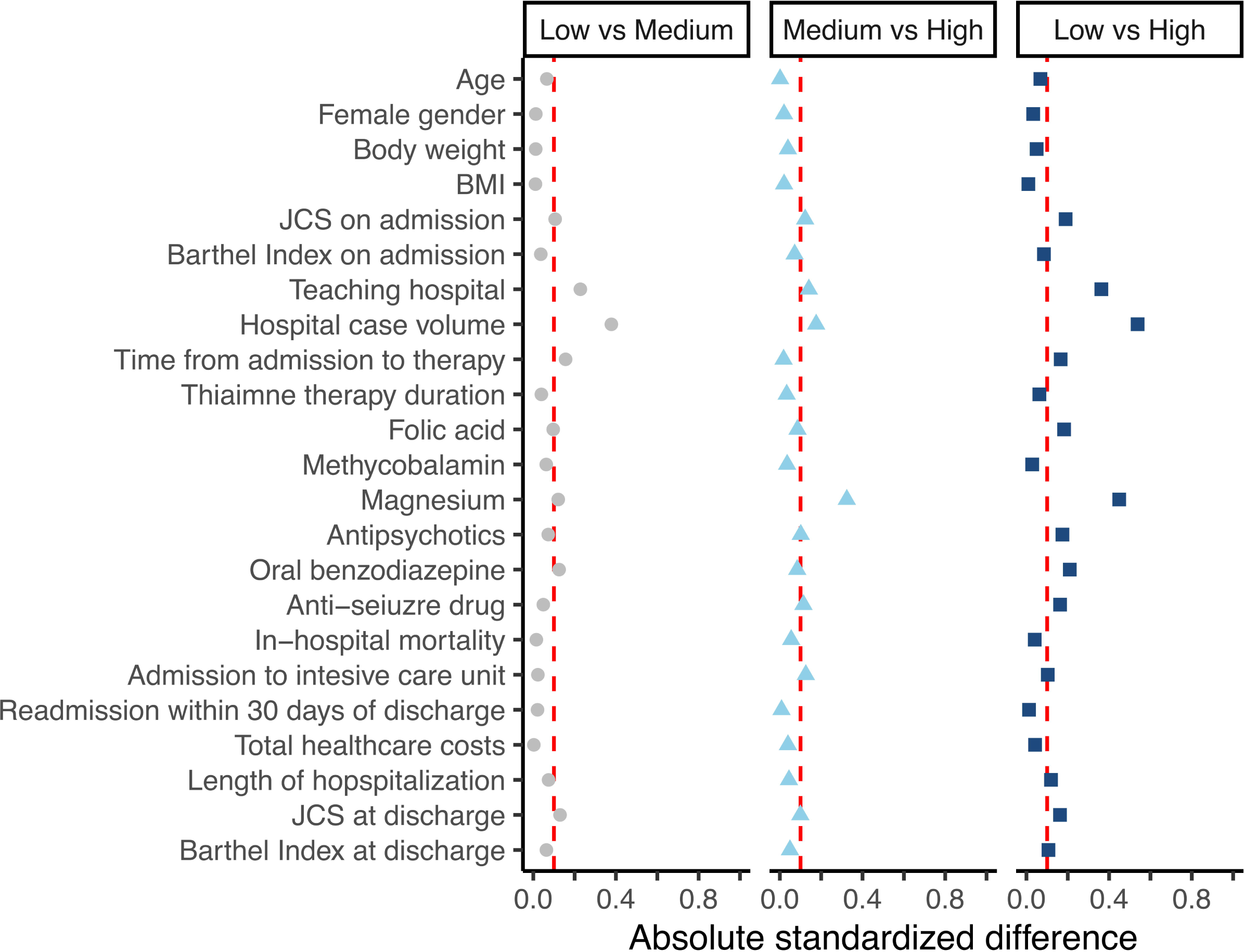
Love plot depicting absolute standardized mean differences (ASDs) across the three initial thiamine dose categories: low (≤300 mg/day), medium (301–900 mg), and high (>900 mg) Dots indicate ASDs for each variable. An ASD threshold of >0.1 denotes a meaningful difference.

**Table 4.** Comparison of the baseline characteristics, treatment patterns, and clinical outcomes stratified by the second-day dose of intravenous thiamine (low, medium, and high) among patients with Wernicke encephalopathy.

|  | Low dose | Medium dose | High dose | Max ASD |
| --- | --- | --- | --- | --- |
|  | ≤300 mg/day | 301–900 mg/day | >900 mg/day |  |
|  | (n = 4329) | (n = 1095) | (n = 2432) |  |
| Age, years, median [IQR] | 66 [56, 74] | 65 [55, 74] | 65 [55, 74] | 0.07 |
| Sex, female | 950 (21.9) | 246 (22.5) | 567 (23.3) | 0.03 |
| Body weight, kg, median [IQR] | 51.2 [44.6, 60.0] | 52.0 [45.0, 60.0] | 51.5 [45.0, 60.0] | 0.05 |
| BMI, kg/m<sup>2</sup>, median [IQR] | 19.5 [17.1, 22.2] | 19.4 [17.3, 22.0] | 19.5 [17.2, 22.1] | 0.02 |
| JCS on admission |  |  |  | 0.19* |
| 0 (Alert) | 1450 (33.5) | 317 (28.9) | 624 (25.7) |  |
| 1–3 (Confused) | 1933 (44.7) | 523 (47.8) | 1254 (51.6) |  |
| 10–30 (Somnolent) | 569 (13.2) | 164 (15.0) | 304 (12.5) |  |
| 100–300 (Comatose) | 343 (7.9) | 89 (8.1) | 241 (9.9) |  |
| Missing | 34 (0.8) | 2 (0.2) | 9 (0.4) |  |
| Barthel Index on admission |  |  |  | 0.09 |
| 100 (Independent) | 544 (12.6) | 135 (12.3) | 253 (10.4) |  |
| 90–95 (Slightly dependent) | 93 (2.1) | 25 (2.3) | 46 (1.9) |  |
| 65–85 (Moderately dependent) | 242 (5.6) | 56 (5.1) | 135 (5.6) |  |
| 0–60 (Severely dependent) | 2665 (61.6) | 701 (64.0) | 1573 (64.7) |  |
| Missing | 785 (18.1) | 178 (16.3) | 425 (17.5) |  |
| Teaching hospital | 3565 (84.5) | 1005 (91.8) | 2316 (95.2) | 0.36* |
| Hospital case volume | 3 [2, 5] | 4 [2, 7] | 5 [2, 8] | 0.54* |
| Intravenous thiamine |  |  |  |  |
| Time from admission to therapy, days, median [IQR] | 0 [0, 1] | 0 [0, 0] | 0 [0, 0] | 0.17* |
| Treatment duration, days, median [IQR] | 6 [3, 10] | 7 [4, 9] | 7 [5, 8] | 0.06 |
| Folic acid† | 971 (22.4) | 291 (26.6) | 740 (30.4) | 0.18* |
| Methylcobalamin† | 1916 (44.3) | 519 (47.4) | 1110 (45.6) | 0.06 |
| Magnesium† | 1187 (27.4) | 361 (33.0) | 1184 (48.7) | 0.45* |
| Antipsychotics† | 1575 (36.4) | 437 (39.9) | 1092 (44.9) | 0.17* |
| Oral benzodiazepine† | 1803 (41.6) | 524 (47.9) | 1266 (52.1) | 0.21* |
| Anti-seizure drug† | 1221 (28.2) | 333 (30.4) | 870 (35.8) | 0.16* |
| In-hospital mortality | 173 (4.0) | 47 (4.3) | 79 (3.2) | 0.08 |
| Admission to the intensive care unit | 346 (8.0) | 81 (7.4) | 268 (11.0) | 0.12* |
| Readmission within 30 days of discharge | 138 (3.2) | 39 (3.6) | 83 (3.4) | 0.05 |
| Total healthcare costs, USD, median [IQR] | 8311 [4640, 13926] | 8704 [5089, 14007] | 8638 [5395, 13695] | 0.07 |
| Length of hospitalization, days, median [IQR] | 23 [11, 43] | 22 [11, 40] | 22 [12, 39] | 0.12* |
| JCS at discharge |  |  |  | 0.16* |
| 0 (Alert) | 3004 (69.4) | 712 (65.0) | 1531 (63.0) |  |
| 1–3 (Confused) | 1065 (24.6) | 307 (28.0) | 772 (31.7) |  |
| 10–30 (Somnolent) | 59 (1.4) | 14 (1.3) | 32 (1.3) |  |
| 100–300 (Comatose) | 16 (0.4) | 13 (1.2) | 14 (0.6) |  |
| Missing | 185 (4.3) | 49 (4.5) | 83 (3.4) |  |
| Barthel Index at discharge |  |  |  | 0.11* |
| 100 (Independent) | 1458 (33.7) | 348 (31.8) | 741 (30.5) |  |
| 90–95 (Slightly dependent) | 243 (5.6) | 59 (5.4) | 118 (4.9) |  |
| 65–85 (Moderate dependent) | 454 (10.5) | 108 (9.9) | 247 (10.2) |  |
| 0–60 (Severe dependent) | 1565 (36.2) | 425 (38.8) | 989 (40.7) |  |
| Missing | 609 (14.1) | 155 (14.2) | 337 (13.9) |  |

IQR, interquartile range; BMI, body mass index; JCS, Japan Coma Scale
Values represent no. (%), except when indicated otherwise.
\*Maximum absolute standardized difference (ASD) >0.1, indicating a meaningful difference.
†Prescribed at least once during hospitalization.

## 4. Discussion

This nationwide Japanese study described intravenous thiamine treatment patterns among patients hospitalized with Wernicke encephalopathy and their clinical outcomes, including in-hospital mortality and functional status at discharge. We found a marked increase in the use of initial high-dose thiamine therapy (>900 mg/day), although approximately half of the patients initially received low-dose treatment (≤300 mg/day) over the study period. More than half of the patients were discharged with impairment in activities of daily living (Barthel Index <90). These findings provide descriptive nationwide data on contemporary thiamine treatment and short-term outcomes in patients with Wernicke encephalopathy.

The annual incidence of Wernicke encephalopathy in fiscal year 2023 was estimated at 1.28 per 100,000 person-years, which is comparable to the rate of 1.03 per 100,000 person-years reported in Spain (2022) [11]. Although another nationwide study conducted in Switzerland (2012–2020) reported a higher annual incidence of 5.82 per 100,000 person-years, its primary analysis included diagnoses in any position. When restricted to patients with a primary or secondary diagnosis at discharge, the incidence in that study was 1.68 per 100,000 person-years, which approximated our estimate [10]. Furthermore, the age distribution, with the largest proportion of patients in the 60–69-year age group, and the predominance of male patients (77.9%) were broadly consistent with those reported in these studies: a mean age of 58.2 years and 75.3% male patients in the Spanish study (2016–2022), and a prominent peak in the 60–65-year age group with 70.8% male patients in the Swiss study (2012–2020). Overall, these findings suggest that the demographic profiles of patients with Wernicke encephalopathy in Japan closely resemble those of European populations.

Following the 2010 European Federation of Neurological Societies guidelines and major review articles published around that time [1–3], a single-center study conducted in France (2008–2018) reported that 1000 mg/day was the most common dose of thiamine, administered to 55.3% of 56 patients [9]. Conversely, another single-center study conducted in Canada (2014–2015) reported that only 5.4% of 17 patients received 1000 mg/day, with 100 mg/day and 500 mg/day being the most frequent doses [8]. Our study showed a significant increasing trend in the use of high-dose regimens (>900 mg/day), as well as a wide variety of regimens and a bimodal distribution of the initial thiamine dose, with peaks at 100 mg/day and 1500 mg/day. The increasing implementation of high-dose therapy, particularly the prominent peak at 1500 mg/day, suggests that recommendations from the guidelines and review articles may have a substantial influence on real-world management in Japan. However, these recommendations are based on limited evidence, and the broad variety of regimens may reflect the limited evidence regarding the optimal thiamine dose. Although a recent single-center, randomized controlled trial compared three categories of initial doses (100 mg, 300 mg, and 500 mg three times daily) and found no significant differences in the improvement of cognitive function and neurological symptoms, the study was hindered by major methodological limitations, such as failure to reach the target sample size and randomization after treatment initiation [23]. Consequently, robust comparative effectiveness research investigating the optimal thiamine dose and associated clinical outcomes is urgently needed.

This study found a relatively low in-hospital mortality rate of 3.8%, which is consistent with previous European studies reporting rates of approximately 5% [7, 10, 11]. However, 55.5% of patients had impairment in activities of daily living (Barthel Index score <90) at discharge, despite prompt intervention, with the median time from admission to the first administration of thiamine being 0 days. A previous Spanish multicenter cohort study demonstrated that 70% of patients with Wernicke encephalopathy were discharged with persistent neurological signs or symptoms [7]. In contrast, another single-center study reported a lower rate of patients experiencing persistent symptoms at 17.6% [24]. However, both of these previous studies lacked standardized functional assessments. Our findings indicate that Wernicke encephalopathy frequently results in severe functional impairment that extends beyond the classic presentation of Korsakoff syndrome, characterized by amnesia and confabulation [1]. This unfavorable functional status, despite immediate thiamine treatment, may be attributable to irreversible structural brain damage existing before hospital admission. Therefore, to improve overall clinical outcomes, there is an urgent need for primary prevention strategies for Wernicke encephalopathy, such as prophylactic thiamine supplementation for high-risk patients [3], in conjunction with further research on the optimal treatment strategy.

In the exploratory analysis, the high-dose thiamine group was associated with a higher proportion of altered mental status on admission, treatment at teaching hospitals and at hospitals with a higher Wernicke encephalopathy case volumes,early thiamine administration, and the use of other concurrent medications. Clinical outcomes such as in-hospital mortality showed no notable difference whereas functional outcomes, including the JCS and Barthel Index scores at discharge, tended to be worse in the high-dose group. These findings suggest that the severity of clinical presentations may lead clinicians to select high-dose therapy, and that teaching hospitals and high-volume hospitals tend to align with the guidelines. Therefore, these outcomes should be interpreted cautiously due to confounding by indication, and further research with adjustment for the baseline characteristics is needed to investigate the true association between thiamine dose and outcomes.

This study has some limitations. First, the estimation of national incidence relied on the assumption that the hospitals included in the DPC database were representative of all acute-care hospitals in Japan. Second, precise information regarding the underlying etiology of Wernicke encephalopathy (alcohol or non-alcohol backgrounds) was not available because the sensitivity of comorbidity diagnoses in the DPC database is generally low [13]. Third, diagnostic misclassification or underestimation may have occurred because the study was based on claims data. Fourth, detailed neurological signs and findings were unavailable. Fifth, although the database allowed us to identify drug prescriptions, it did not capture whether the prescribed drugs were actually administered. Sixth, the time from symptom onset to admission was not available, which may have affected the clinical outcomes. Seventh, data on the pre-admission JCS and Barthel Index scores were unavailable.

Consequently, it was difficult to determine whether the unfavorable JCS and Barthel Index scores at discharge were attributable to Wernicke encephalopathy or pre-existing baseline impairments. Finally, this study could not document the long-term prognosis after discharge.

## 5. Conclusions

The use of high-dose thiamine has increased significantly over the past 13 years. While the in-hospital mortality rate was relatively low, approximately half of the patients with Wernicke encephalopathy were discharged with impairment in activities of daily living. To improve overall outcomes, further research to establish the optimal thiamine treatment strategy and primary prevention measures is urgently needed.

## Statements and Declarations

### Ethics approval

This study was approved by the Institutional Review Board of the University of Tokyo [approval number: 3501– (5)] and conducted in accordance with the Declaration of Helsinki.

### Consent

The requirement for informed consent was waived because of the anonymous nature of the database.

### Data availability

The data that support the findings of this study are not publicly available due to data use agreements with the participating hospitals and data providers.

### Author contributions

Conceptualization: N.Y. and Y.K.; methodology: N. Y. and Y.K.; software: N.Y., Y.K., H.M., and K.F.; formal analysis: N.Y. and Y.K.; writing the original draft: N.Y.; writing, review, and editing: N.Y., Y.K., H.M., K.F., and H.Y.; supervision: Y.K. and H.Y. All authors have participated in the interpretation of the results and writing of the report, and agreed to the published version of this manuscript.

## Acknowledgements

This work was supported by grants conferred by the Ministry of Health, Labor and Welfare, Japan (23AA2003 and 24AA2006).

